# Right Ventricular Dysfunction: An Overlooked Predictor of Sudden Cardiac Death and Arrhythmic Events – A Meta-Analysis

**DOI:** 10.1101/2025.07.12.25330929

**Authors:** Toshinori Chiba, Amelie Beblo, Nora Wainstejn, Julia Lueg, Robert Hättasch, Felix Hohendanner, Verena Tscholl, Nikolaos Dagres, Gerhard Hindricks, Wilhelm Haverkamp

## Abstract

**Background and Aim:** Right ventricular (RV) dysfunction has emerged as a potential predictor of adverse cardiac outcomes, including sudden cardiac death (SCD) and severe ventricular arrhythmias (VA). To clarify the role of RV dysfunction in arrhythmic risk assessment, we conducted a systematic review and meta-analysis to evaluate its prognostic value for SCD and VA.

**Methods:** A systematic search of PubMed, Embase, and Web of Science was conducted from inception to February 2025. The primary endpoint was a composite of SCD or severe VA, including ventricular tachycardia, ventricular fibrillation, or appropriate implantable cardioverter-defibrillator therapy. RV dysfunction was defined as impaired RV fractional area change, RV ejection fraction, or RV strain. A random-effects model was used to calculate pooled risk ratios (RRs) and mean differences (MDs) with 95% confidence intervals (CIs).

**Results:** Of 1,296 identified articles, seven studies comprising 1,475 patients met inclusion criteria. RV dysfunction was present in 541 patients. The mean follow-up duration was 2.9 years. RV dysfunction was significantly associated with an increased risk of SCD and VA (RR: 3.73; 95% CI: 2.43–5.72; p < 0.01). Secondary analysis showed that patients who experienced SCD or VA had significantly lower RVFAC compared to those without events (MD: −5.67; 95% CI: −8.73 to −2.60; p < 0.01), whereas LVEF did not differ significantly between groups (MD: −0.59; 95% CI: −4.10 to 2.92; p = 0.74).

**Conclusions:** RV dysfunction is associated with a increased risk of SCD and VA, indicating that RV parameters may hold prognostic value in arrhythmic risk stratification.

## Introduction

Sudden cardiac death (SCD) remains a major public health concern, with an estimated global incidence ranging from 30 to 100 per 10,000 individuals annually [1, 2]. Despite advances in emergency response systems—such as automated external defibrillators and increased rates of bystander cardiopulmonary resuscitation—ventricular arrhythmias (VAs), particularly ventricular tachycardia (VT) and ventricular fibrillation (VF), remain the predominant causes of out-of-hospital cardiac arrests [3].

Left ventricular (LV) dysfunction, especially a reduced LV ejection fraction (LVEF), is a well-established risk factor for SCD and underpins current guideline recommendations for implantable cardioverter-defibrillator (ICD) implantation in primary prevention. Patients with an LVEF ≤35% are considered at high risk and are eligible for ICD therapy, which has been shown to reduce mortality by preventing life-threatening arrhythmias [4]. Accordingly, both American and European guidelines (AHA/ACC/HRS and ESC) emphasize LVEF as a central criterion for ICD implantation [5, 6].

However, growing evidence suggests that reliance on LVEF alone may be inadequate for comprehensive risk stratification. Notably, Stecker et al. reported that only approximately one-third of SCD cases occur in patients with an LVEF ≤35%, while nearly half occur in individuals with preserved LV function [7]. This discrepancy underscores the need for additional, more sensitive predictors of arrhythmic risk. Emerging data point to the prognostic significance of right ventricular (RV) dysfunction in this context. RV impairment has been associated with adverse outcomes in a range of cardiac conditions, including heart failure progression and increased susceptibility to VAs. Several studies, including those by Pandat et al. and our own group, have demonstrated that reduced right ventricular fractional area change (RVFAC) is independently associated with an increased risk of SCD or appropriate ICD therapy [8, 9]. Moreover, Elming et al. observed that among patients with LV dysfunction, coexisting RV dysfunction was linked to a greater survival benefit from ICD implantation, further suggesting a role for RV function in refining risk stratification [10].

In light of these findings, we conducted a systematic review and meta-analysis to evaluate the association between RV dysfunction and the risk of SCD and VA. We also sought to compare the prognostic value of RV function with LVEF to assess whether RV parameters may offer incremental benefit in arrhythmic risk prediction.

## Methods

This meta-analysis was conducted in accordance with the Preferred Reporting Items for Systematic Reviews and Meta-Analyses (PRISMA) guidelines [11]. The study protocol was prospectively registered in the PROSPERO database (registration number: CRD420251045088).

### Eligibility Criteria

Studies were eligible for inclusion if they met the following criteria: (1) observational cohort studies (prospective or retrospective) or randomized controlled trials; (2) investigated the association between RV dysfunction and SCD or VA events; (3) provided extractable data for risk estimation (e.g., number of events, hazard ratios, odds ratios, or risk ratios), included a minimum follow-up period of at least 12 months, and assessed RV function using echocardiography or cardiac magnetic resonance imaging (MRI).

We excluded case reports, editorials, letters, conference abstracts, systematic reviews, meta-analyses, and studies without extractable outcome data. A secondary objective was to compare the prognostic utility of RV dysfunction with that of LVEF in predicting SCD or VA.

### Data Sources and Search Strategy

A comprehensive literature search was performed using PubMed, Embase, and Web of Science from database inception to February 2024. Search terms included: (“right ventricular dysfunction” OR “right ventricular function”) AND (”sudden death” OR “ventricular arrhythmia” OR “ICD” OR “implantable cardioverter defibrillator”). Two reviewers independently screened all titles and abstracts, followed by full-text review. Disagreements were resolved by discussion or consultation with a third reviewer. Reference lists of all eligible articles were manually searched for additional studies.

### Data Extraction

Data were extracted using a standardized form and included: study characteristics (first author, publication year, study design); patient demographics (sample size, age, sex distribution, comorbidities, and clinical background), RV functional parameters (RVFAC, RV ejection fraction (RVEF), RV strain), primary outcomes (SCD, VA, and appropriate ICD therapy - shock or anti-tachycardia pacing), and; and follow-up duration.

### Endpoint Definitions

The primary endpoint was a composite of SCD or VA, defined as: SCD or aborted SCD (documented VT/VF, resuscitated cardiac arrest), and appropriate ICD therapy (shock or anti-tachycardia pacing). These endpoints were collectively referred to as “SCD or VA events” for analysis.

### Statistical Analysis

Statistical analyses were performed using R software version 4.4.3 (R Foundation for Statistical Computing, Vienna, Austria) and followed the Cochrane Handbook for Systematic Reviews of Interventions [12]. Dichotomous outcomes were pooled using random-effects models to calculate pooled risk ratios (RRs) and hazard ratios (HRs) with 95% confidence intervals (CIs). Continuous outcomes were analyzed using inverse-variance random-effects models to estimate mean differences (MDs).

If necessary, missing event numbers were estimated using the method of Tierney et al., which allows for calculation of log HRs from published data [13]. For studies reporting medians and interquartile ranges, means and standard deviations were approximated using the method of Luo et al [14].

Heterogeneity was assessed using Cochran’s Q test and the I² statistic. Sensitivity analyses were conducted to examine the impact of data estimation methods. A secondary analysis was performed to compare the prognostic relevance of RV versus LV function.

Study quality was evaluated using the Newcastle–Ottawa Scale [15,16]. Publication bias was assessed using funnel plots and Egger’s regression test, with p-values < 0.05 indicating potential bias. A p-value < 0.05 was considered statistically significant throughout.

## Results

### Study Selection

A total of 1,296 records were identified through database searches: 462 from PubMed, 464 from Embase, and 370 from Web of Science. After removal of 446 duplicates, 850 records remained for title and abstract screening. Of these, 806 were excluded as irrelevant to the study objective. The full texts of 44 articles were assessed for eligibility, and 7 studies [8,9,10,17–20] met the inclusion criteria for the primary meta-analysis. One additional study [21] was included solely for secondary analysis. The study selection process is illustrated in the PRISMA flowchart (Figure 1).

**Figure 1.**
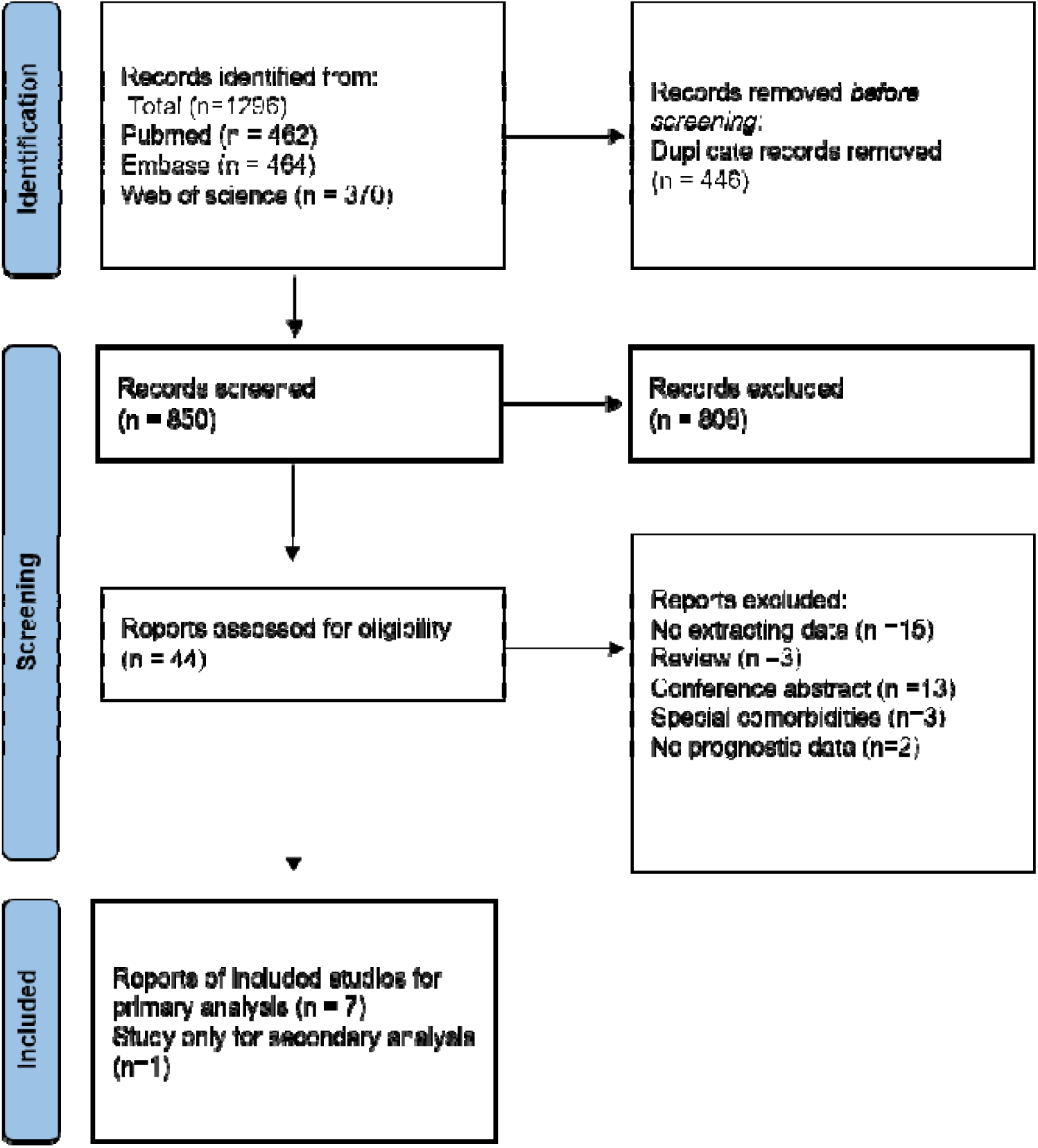
The flowchart illustrating selection process.

### Study Characteristics

Table 1 summarizes the characteristics of the included studies. Studies 1 through 7 were included in the primary meta-analysis. Study 8 was used exclusively for the secondary analysis, as it stratified patients based on appropriate ICD shocks, making extraction of RV dysfunction-specific outcome data infeasible.

**Table 1.**
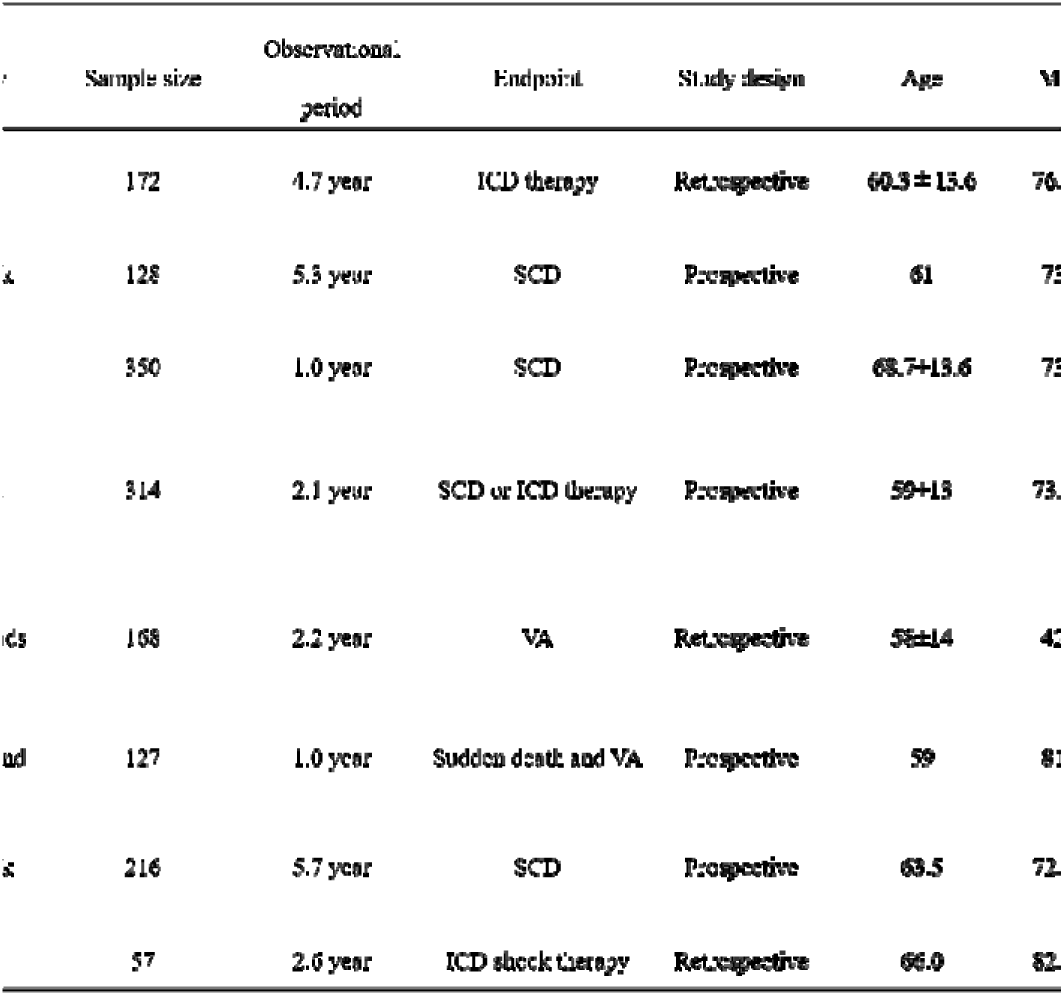
Study characteristics. ICD, implantable cardioverter-defibrillator; LVEF, left ventricular ejection fraction; RVFAC, right ventricular fractional area change, RVEF, right ventricular ejection fraction; RV-FWS, right ventricular free wall strain; SCD, sudden cardiac death; VA, ventricular arrhythmia

A total of 1,475 patients were included, of whom 541 had RV dysfunction. The mean follow-up duration across studies was 2.9 years. RV function was assessed using RVFAC in two studies and RVEF in four studies; one study employed RV free wall strain. Five of the seven primary studies had a prospective design. Study populations varied: one was community-based (a registry of SCD), and four included patients with ischemic or non-ischemic cardiomyopathy.

Quality assessment using the Newcastle–Ottawa Scale (Table 2) showed that studies scored between 6 and 9 points, with a mean score of 7.9. Most studies were classified as high quality; only one was considered of moderate quality. Egger’s regression test indicated no significant publication bias (p = 0.58).

**Table 2.**
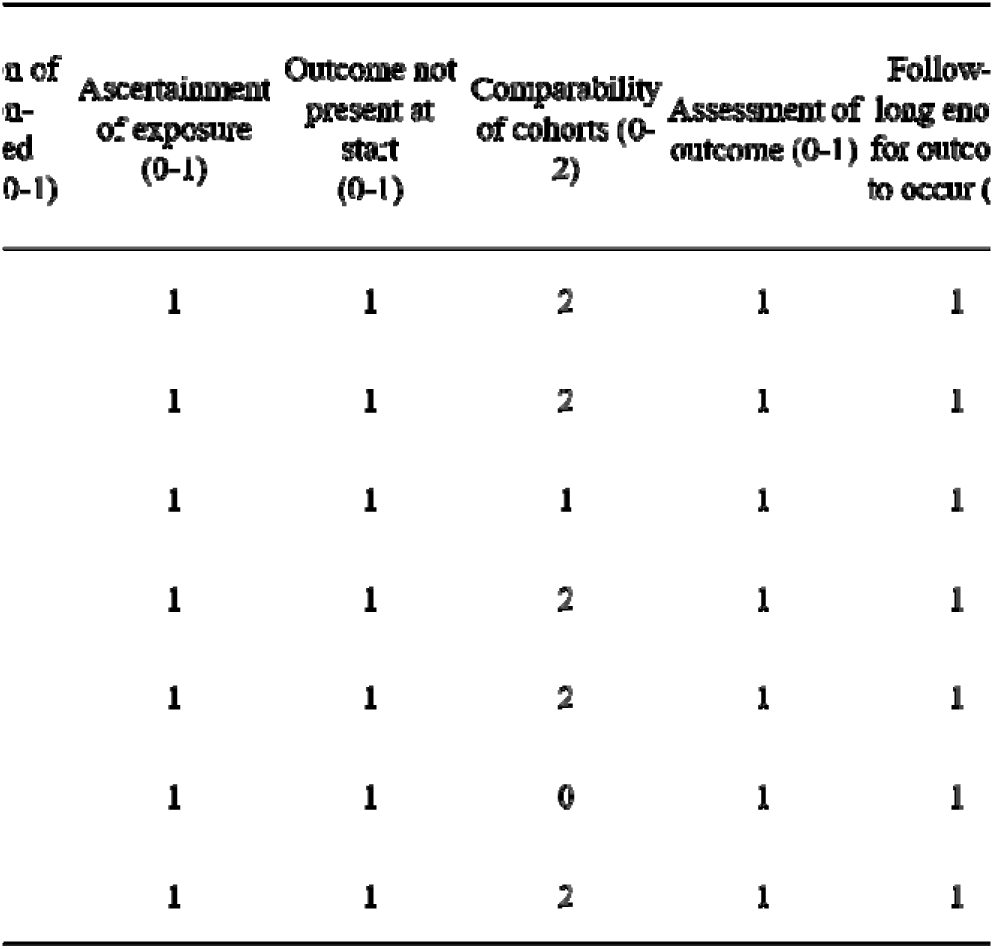
Quality Assessment Using the Newcastle Ottawa Scale.

### RV Dysfunction and Risk of Sudden Cardiac Death or Ventricular Arrhythmia

During follow-up, 223 patients experienced the composite outcome of SCD or VA. RV dysfunction was significantly associated with a higher risk of SCD or VA (RR: 3.73; 95% CI: 2.43–5.72; p<0.01), with moderate heterogeneity (I² = 43%, p = 0.11), as shown in Figure 2. In a subgroup analysis restricted to studies enrolling patients with ischemic or non-ischemic cardiomyopathy, the association remained significant (RR: 2.93; 95% CI: 1.53–5.61; I² = 42%, p = 0.16) (Figure 3).

**Figure 2.**
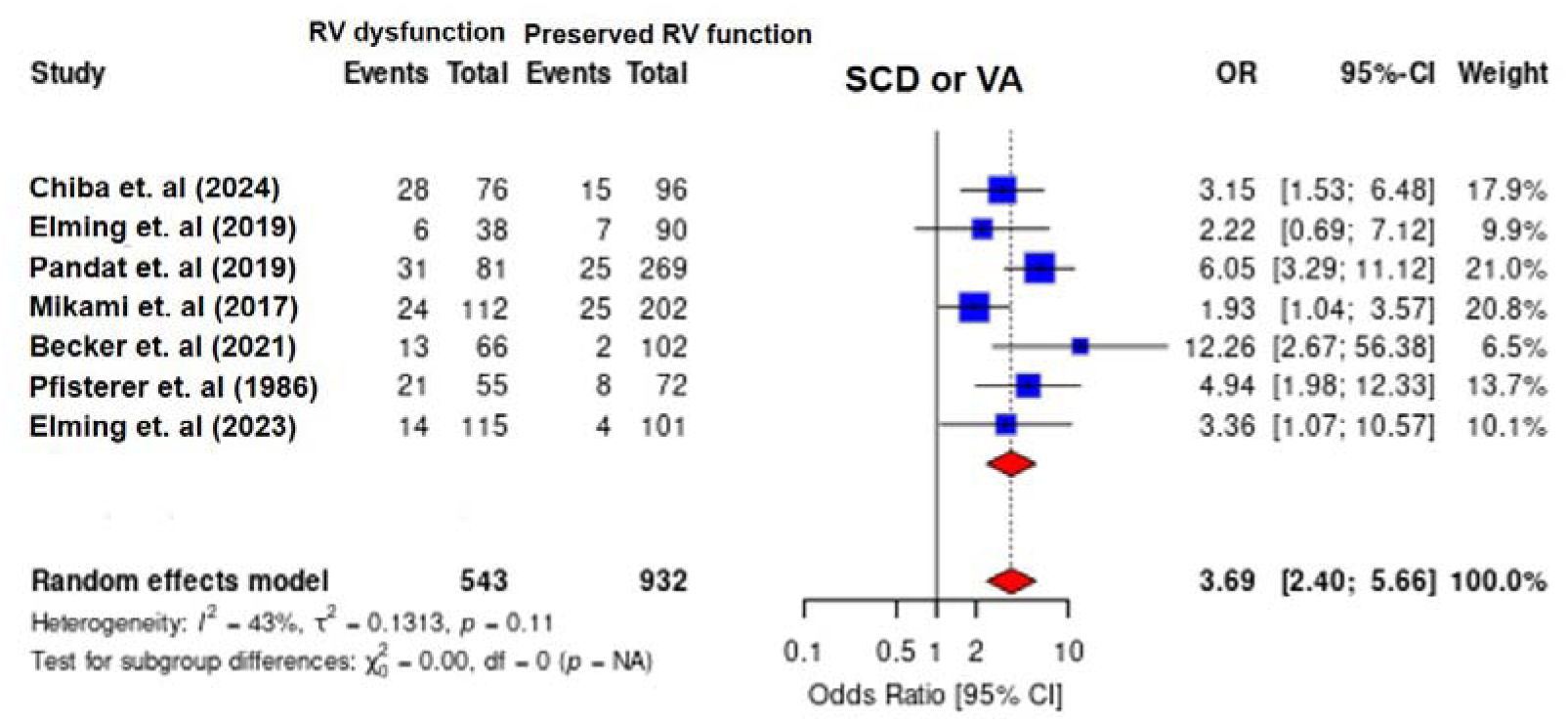
Forest plot illustrating the association between right ventricular dysfunction and the risk of sudden cardiac death or ventricular arrhythmia. RV dysfunction was significantly associated with an increased risk of sudden cardiac death or ventricular arrhythmia, with a pooled odds ratio (OR) of 3.69 (95% CI: 2.40–5.66) and moderate heterogeneity (I² = 43%). RV, right ventricular; SCD, sudden cardiac death; VA, ventricular arrhythmia

**Figure 3.**
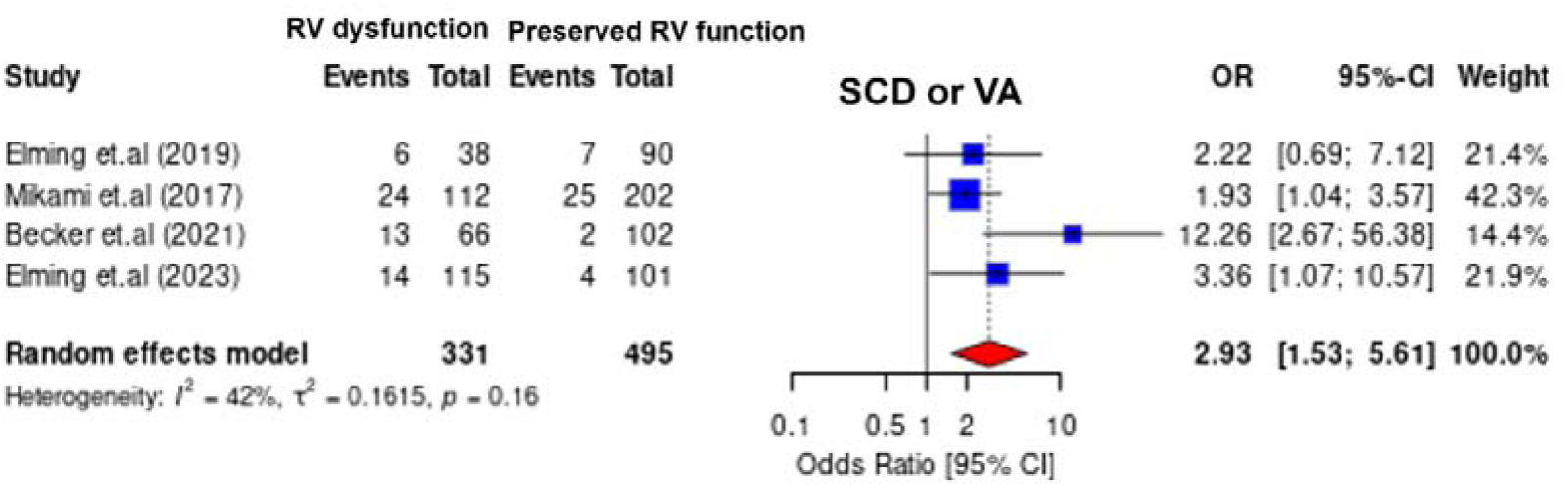
Forest plot illustrating the association between right ventricular dysfunction and the risk of sudden cardiac death or ventricular arrhythmia among studies limited to patients with ischemic or non-ischemic cardiomyopathy. In a subgroup analysis including four studies, RV dysfunction remained significantly associated with an increased risk of SCD or VA (OR: 2.93; 95% CI: 1.53–5.61), with moderate heterogeneity (I² = 42%). RV, right ventricular; SCD, sudden cardiac death; VA, ventricular arrhythmia

### Comparison of LV and RV Function in Predicting SCD and VA

Three studies (Chiba et al. 2024, Pandat et al. 2019, Malasana et al. 2011) provided both LV and RV function data in patients who experienced SCD or VA. Two of these studies included patients with ICDs implanted for primary or secondary prevention; the third was a community-based registry.

Using variance-based random-effects models, we compared the prognostic value of LVEF and RVFAC. LVEF did not significantly differ between patients with and without SCD or VA (MD: −0.59; 95% CI: −4.10 to 2.92; p = 0.74). In contrast, RVFAC was significantly lower in patients with SCD or VA (MD: −5.67; 95% CI: −8.73 to −2.60; p < 0.01) (Figure 4).

**Figure 4.**
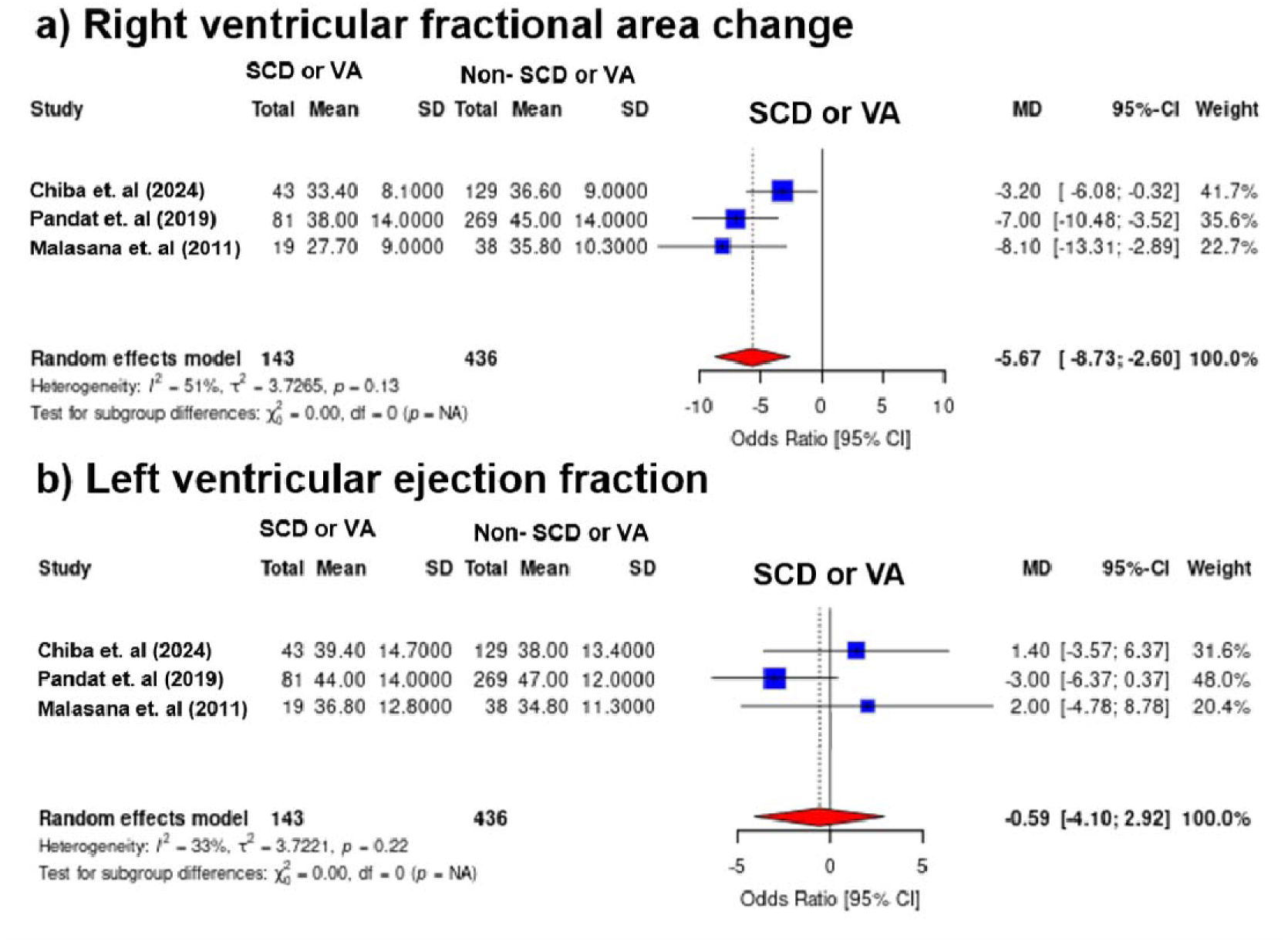
Forest plots illustrating comparisons of right ventricular (RV) fractional area change and left ventricular ejection fraction (LVEF) between patients with and without sudden cardiac death (SCD) or ventricular arrhythmia (VA). (a) Sudden cardiac death and ventricular arrhythmia was related to low RVFAC. (b) In contrast, there was no significant difference in LVEF between the two groups. Patients with SCD or VA had significantly worse RV dysfunction compared to those without events, whereas no significant difference was observed in left ventricular ejection fraction between the two groups. SCD, sudden cardiac death; VA, ventricular arrhythmia

### Sensitivity Analysis

Sensitivity analyses showed that the results were robust across all leave-one-out analyses. Notably, exclusion of Mikami et al. (2017) reduced heterogeneity substantially (I² from ∼49% to 4%), indicating this study was a major source of heterogeneity without altering the overall effect estimate.

## Discussion

In this meta-analysis of seven studies, we evaluated the association between RV function and the risk of SCD and severe VA. We also compared the prognostic value of RV function with that of LV function. To our knowledge, this is the first meta-analysis focusing specifically on RV function in relation to SCD and VA, offering new insights into risk stratification and potential indications for ICD therapy.

Our findings indicate: (1) RV dysfunction is significantly associated with an increased risk of SCD and VA. (2) In both ICD cohorts and non-selective populations, RV dysfunction—but not LV dysfunction—was predictive of SCD and VA.

### RV Dysfunction and Prognosis

RV function has emerged as a critical determinant of prognosis in various cardiovascular conditions. Previous meta-analyses have demonstrated that RV dysfunction, assessed by cardiac MRI or echocardiography, predicts higher rates of mortality and hospitalization in patients with heart failure [22,23]. Notably, Gorter et al. showed that RV dysfunction (based on tricuspid annular plane systolic excursion and RVFAC) was also associated with worse outcomes in patients with heart failure with preserved ejection fraction (HFpEF) [24].

These associations extend beyond traditional heart failure. RV dysfunction has been linked to adverse outcomes in myocarditis—including death, hospitalization, and arrhythmia [25]—and in pulmonary hypertension, where it is associated with increased risk of SCD and VA independently of LV function [26]. Similarly, in cardiac sarcoidosis, RV dysfunction has been tied to both VAs and SCD [27]. Collectively, these findings highlight the prognostic relevance of RV dysfunction across a range of cardiovascular diseases.

### Mechanistic Importance of RV Function

The pathophysiological mechanisms linking RV dysfunction to adverse outcomes are increasingly recognized. While LV function has historically been emphasized, RV performance plays a key role in maintaining cardiac output by providing preload to the LV. RV dilation and interventricular dependence can impair LV filling [28], and combined RV systolic and biventricular diastolic dysfunction may reduce both cardiac output and coronary perfusion [29].

In HFpEF, approximately one-third of patients exhibit RV dysfunction [30]. These patients often show elevated pulmonary artery pressures, and progressive RVFAC decline has been linked to worsening pulmonary hypertension [30]. Thus, RV dysfunction may critically affect pulmonary and systemic circulations, reinforcing its prognostic significance.

### Clinical Implications for SCD and VA Risk Stratification

This meta-analysis demonstrates that RV dysfunction is strongly associated with SCD and VA. Even in subgroups restricted to ischemic or non-ischemic cardiomyopathy, this association remained robust. These findings suggest that RV dysfunction could serve as a valuable risk marker for identifying patients at high risk of life-threatening arrhythmias—particularly those who may benefit from ICD therapy.

Importantly, our secondary analysis found that RV function was more predictive of SCD and VA than LV function, even in ICD recipients (both primary and secondary prevention) and non-selective populations. Xing et al. also found that low RVEF was associated with fatal arrhythmic events in patients without structural heart disease [31], further supporting the value of RV function as an independent risk factor.

### Limitations

This study has several limitations. First, the definitions and assessments of RV dysfunction varied across the included studies, with different parameters and cutoff values used. This heterogeneity could affect the comparability of the results. Second, baseline patient characteristics varied among studies, which may introduce variability in the findings. The small number of studies available limited our ability to conduct more detailed subgroup analyses, and we focused on ischemic and non-ischemic cardiomyopathy as a result. Third, we were unable to separately analyze patients with ICDs based on primary versus secondary prevention indications, which could have provided additional insights.

### Conclusion

Our meta-analysis demonstrates that RV dysfunction is significantly associated with an increased risk of SCD and VA. Furthermore, RV dysfunction appears to be a stronger prognostic indicator than LVEF in predicting these outcomes. These findings underscore the importance of incorporating RV function into risk stratification and clinical decision-making for patients at risk of SCD. However, given the variability in RV dysfunction definitions and the small number of studies included, further large-scale, well-designed studies are necessary to validate these findings, establish standardized assessment criteria for RV function, and better define its role in clinical practice, particularly for risk stratification and ICD therapy.

## Data Availability

All data produced in the present study are available upon reasonable request to the authors

